# Early screening of colorectal cancer using feature engineering with artificial intelligence-enhanced analysis of nanoscale chromatin modifications

**DOI:** 10.1101/2023.10.30.23297790

**Authors:** Andrew Chang, Sravya Prabhala, Ali Daneshkhah, Jianan Lin, Hariharan Subramanian, Hemant Kumar Roy, Vadim Backman

## Abstract

Colonoscopy is accurate but inefficient for colorectal cancer (CRC) prevention due to the low (~7-8%) prevalence of target lesions, advanced adenomas. We leveraged rectal mucosa to identify patients who harbor CRC field carcinogenesis by evaluating chromatin 3D architecture. Supranucleosomal disordered chromatin chains (~5-20 nm, ~1 kbp) fold into chromatin packing domains (~100-200 nm, ~100–1,000 kbp). In turn, the fractal-like conformation of DNA within chromatin domains and the folding of the genome into packing domains has been shown to influence multiple facets of gene transcription, including the transcriptional plasticity of cancer cells. We deployed an optical spectroscopic nanosensing technique, chromatin-sensitive partial wave spectroscopic microscopy (csPWS), to evaluate the packing density scaling D of the chromatin chain conformation within packing domains from rectal mucosa in 256 patients with varying degrees of progression to colorectal cancer. We found average packing scaling D of chromatin domains was elevated in tumor cells, histologically normal-appearing cells 4 cm proximal to the tumor, and histologically normal-appearing rectal mucosa compared to cells from control patients (p<0.001). Nuclear D had a robust correlation with the model of 5-year risk of CRC with r2=0.94. Furthermore, rectal D was evaluated as a screening biomarker for patients with advanced adenomas presenting an AUC of 0.85 and 85% sensitivity and specificity. Artificial Intelligence (AI)-enhanced csPWS improved diagnostic performance with AUC=0.90. Considering the low sensitivity of existing CRC tests, including liquid biopsies, to early-stage cancers our work highlights the potential of chromatin biomarkers of field carcinogenesis in detecting early, significant precancerous colon lesions.

## Introduction

Colorectal cancer (CRC) is the third-most diagnosed cancer in males and second in females with over 52,000 annual US fatalities[1]. Improvements in the detection of CRC at earlier stages and more effective primary and adjuvant treatment options have resulted in decreased mortality rates due to CRC over the past 30 years in the United States and other Western countries[2, 3]. Colonoscopy is the current gold standard screening modality, but attempting to perform colonoscopy on the entire average-risk population is inefficient, as only 7-8% have advanced adenomas. The direct visualization of adenomatous polyps within the field of view of the endoscope offers excellent sensitivity to treatable, early-stage precancerous lesions and provides the opportunity to remove advanced adenomas (stage AA, size > 1cm or > 25% vilous features or high-grade dysplasia) that may later progress into invasive CRC. However, colonoscopy is hampered by patient noncompliance, the inconvenience of bowel preparation, the potential requirement for dietary and medical adjustments, the potential for sedation-related complications, and procedural risks of perforation, major bleeding, and infection [4, 5]. Current efforts to reduce CRC incidence and mortality, particularly for younger adults, are focused on identifying patients who warrant earlier screening through increased public awareness of cancer risk and symptoms and the development of early risk stratification tools with high sensitivity and accessibility[3, 6, 7].

Among the different types of screening techniques are stool-based and blood-based tests. Stool-based testing includes fecal immunochemical test (FIT) and guaiac-based fecal occult blood test (gFOBT), which detects either blood or hemoglobin, and multitarget stool DNA test (sDNA-FIT, Cologuard), which is a molecular assay to test for tumor DNA mutations and methylation markers [8–12]. Stool-based testing has the advantage of noninvasiveness and better patient uptake[13]. Fecal tests have also been shown to decrease CRC incidence, albeit modestly[10]. The sensitivity of FIT for AA is 21-25%[10]. The Cologuard test combines FIT with KRAS mutation and 2 methylation markers with sensitivity of 42% for stage AA but is counterbalanced by lower specificity (and hence more false positives) and cost (~10 times the cost of FIT alone)[14]. Recently, there has been significant interest in liquid biopsy tests which are capable of detecting genetic and epigenetic modifications and fragmentation in circulating tumor DNA (ctDNA)[15, 16]. Companies including Grail, Freenome, Guardant, Delfi, and Thrive have actively developed liquid biopsy tests as a potential cancer screening modality [17–25]. Their initial results demonstrated the capability to detect various cancers, including CRC; however, their sensitivity to early-stage disease dropped precipitously below a clinically acceptable level. The main limitation of such tests is due to the limited amount of DNA released by a tumor into circulation, with smaller lesions secreting less tumor ctDNA (~ 1 ctDNA/ 10 mL of blood)[26–28]. For example, a recent study revealed that ctDNA was detected in 45% of CRC cases, whereas its presence was observed in less than 2.6% of advanced adenoma cases[29]. The considerable heterogeneity in tumor cells complicates the evaluation of DNA fragmentation or specific genetic/epigenetic changes in clinically accepted blood samples using liquid biopsy tests for detecting small lesions. Guardant’s recent ECLIPSE trial showed a drop in performance from overall sensitivity of 83% for CRC to 13% for advanced adenoma[24]. The Shield blood test that utilizes genetic, epigenetic, and proteomics from circulating tumor DNA demonstrated sensitivity of 91% in CRC, 20% in advanced adenoma with a specificity of 92%. Similarly low performance for screening advanced adenomas was observed with Freenome’s recently published AI-EMERGE study (n=664) with an overall sensitivity of 41% and specificity of 90%, which is decreased (sensitivity of 25%) when the size of the advanced adenoma is limited to less than 10 mm[30]. A sensitive, accurate, accessible, and cost-efficient test that is not restricted by lesion size may therefore provide significant clinical value. A successful test design requires three crucial elements: an accessible biomarker source, a biomarker that is sensitive to advanced adenoma, and a modality that enables population-wide screening.

Here we explore field carcinogenesis as an alternative biomarker source. Carcinogenesis involves the complex interplay between environmental exposures and genetic / epigenetic status. Field carcinogenesis is the process by which cells throughout the colonic mucosa accumulate carcinogenic alterations, and due to stochastic events, some of these give rise to a tumor clone. As cells throughout the colonic mucosa harbor these carcinogenic alterations, field carcinogenesis can be utilized as a robust marker to assess the risk of neoplasia for the entire colon[31, 32]. Field carcinogenesis is the underpinning of the clinical practice of surveillance colonoscopy—performing more frequent colonoscopy in patients with a prior adenoma since they are at higher risk of developing new polyps throughout the colon. Flexible sigmoidoscopy allows cancer screening from a more accessible site, and identification of adenomas in the distal colon is associated with a 2.5-fold higher risk of proximal neoplasia[2]. Several studies have shown the efficacy of flexible sigmoidoscopy as a risk stratification tool in cancer prevention and reduced mortality through utilization of field carcinogenesis[33, 34]. Aside from these morphological markers, in the visually normal colonic mucosa rectal mucosa there are myriad cellular, physiological, genomic/proteomic, epigenetic, and molecular events that correlate with concurrent and future neoplasia[35, 36]. Cellular markers of neoplasia include increased proliferation and decreased apoptosis. Physiologically, there is evidence of an early increase in blood supply potentially driven by metabolic changes (Warburg effect). There are multiple genes and proteins altered in the normal colonic mucosa. From an epigenetic perspective, both microRNA and methylation have been shown to be altered[36, 37]. The occurrence of multiple synchronous and metachronous primary neoplastic development, and local recurrence can be well explained by field carcinogenesis[35, 37]. Several studies were conducted on specific epigenetic alterations such as hypermethylation of CpG island by Tahara et. al. and hypomethylation in LINE-1 by Kamiyama et. al. in CRC progression. Along with studies that directly examined gene and epigenetic alterations, other studies demonstrated that chromatin structural changes may also affect silencing of tumor suppressor genes[38]. The dynamic chromatin structure, which modulates gene expression by controlling the accessibility of transcription factors (TF) and RNA polymerases (RNAPs), also holds potential to be utilized as a predictive tool for detection of early-stage cancer.

We explored 3D chromatin structure as a biomarker of colorectal carcinogenesis. Chromatin adopts a complex structure across multiple length scales. At the smallest scale, DNA wraps around histones to form nucleosome complexes colloquially known as “beads on a string.” Nucleosomes and linker DNA then organize into disordered chains with diameters spanning from 5 to 24 nm that typically comprise 200 – 1,000 bp. The chromatin chain is packed at varying volume concentrations to form packing domains (PDs) with an average genomic size of approximately 200 kbp and average physical radius of around 80 nm[39–42]. Within PDs, chromatin follows a scaling relationship between the number of chain monomers (N_f_) and the space it occupies that is well approximated as a power law (N_f_ ∝ r^D^), thus exhibiting a mass fractal-like polymer conformation behavior. Accordingly, conformation of chromatin inside a packing domain can be characterized by chromatin density packing scaling exponent D, which provides insight into the physical nanoarchitecture of chromatin. PDs play a crucial role in transcriptional regulation. Gene transcription tends to occur at the periphery of PDs, and PD structure as well as genomic processes that regulate the emergence, maintenance, and dissipation of PDs have direct implications for the rates of transcriptional reactions and new transcriptional up- or downregulation [40]. The dysregulation of chromatin PDs has been implicated in transcriptional alterations during carcinogenesis. For example, a higher value D of a domain is associated with lower gene connectivity scaling[43, 44] and more frequent long-distance gene loci contacts [43, 45]. Presence of high-D PDs and greater packing domain upregulation have been causally linked with several transcriptional patterns prevalent in cancer cells, including transcriptional divergence (further upregulation of initially upregulated genes with simultaneous suppression of downregulated genes)[43], transcriptional malleability (enhanced rates of new transcriptional upregulation), and transcriptional intercellular heterogeneity (the standard deviation of expression of genes across a cell population). Taken together, these processes enhance the ability of cancer cells to attain new transcriptional states [42]. Neoplastic cells may derive advantages from transcriptional plasticity as they must adapt and acquire new traits in response to different constraints and changes in the microenvironment and host responses [40, 43]. Consequently, chromatin 3D architecture can serve as a marker for the progression of neoplastic changes.

Changes in chromatin domain structure occur at various length scales, ranging from approximately 20 nm to 300 nm [46]. Conventional optical microscopy lacks the ability to differentiate structures smaller than half the wavelength of visible light, which typically ranges from 400 to 750 nm. To overcome this limitation, we have developed an optical spectroscopic statistical nanosensing approach known as csPWS, or chromatin-sensitive partial wave spectroscopic microscopy. csPWS enables calculation of the packing scaling behavior of chromatin PDs within the nucleus, thereby enabling sensitivity to structural changes that are smaller than half the wavelength of visible light at a length scale sensitivity of 23 – 334 nm[40]. This is accomplished by analyzing the spatial variations in the refractive index (RI) through spectroscopic analysis of the interference of scattered light within each diffractional resolution voxel[47, 48]. For a given cell, the output of csPWS microscopy is an image of a nucleus where each pixel represents the packing scaling behavior of chromatin PDs. This image highlights the structural heterogeneity within a coherence volume centered around each pixel. The packing scaling D is estimated by measuring the standard deviation of the spectra generated by the interference of light scattered by the spatial variations of the chromatin density and a reference wave and applying the framework provided in [49]. Our optical statistical nanosensing approach enables a high throughput, robust, and reproducible characterization of chromatin organization and provides valuable insights into its structural properties at the nanoscale.

Prior studies have shown that although intra-domain scaling D is a powerful regulator of transcriptional plasticity, other properties of chromatin 3D structure may play a substantial regulatory or modulating role. Factors including nuclear crowding density, genomic size (Nd) of a domain, domain volume fraction as a function of intranuclear (e.g., peripheral vs interior) location, interdomain interactions, histone modification in and outside of domains, and others may affect chromatin connectivity, accessibility, transcriptional malleability and heterogeneity, and ultimately global patterns of gene expression[40, 42]. These factors influence the chromatin structure and its functional properties within the nucleus. The average nuclear packing scaling D does not fully capture the complexity of dynamic chromatin structural changes. Thus, advanced machine learning and artificial intelligence (AI) deployed on csPWS images of cell nuclei can be utilized to more accurately capture the complexity of these chromatin properties.

In this study, we bridged field carcinogenesis as a biomarker source and chromatin domain dysregulation as the biomarker with recently developed csPWS microscopy to develop and test a new approach to early CRC screening, where cells are obtained by brushing the rectal mucosa, followed by csPWS measurement of their chromatin structure with the resulting data being further analyzed with the help of machine learning. We evaluated chromatin structural alterations within and across PDs within cell nuclei of rectal cells, optimized cell acquisition and analysis, identified and optimized chromatin biomarkers of field carcinogenesis, and tested the diagnostic accuracy of this approach for the identification of patients who harbor pre-cancerous advanced adenomas in the colorectal mucosa. The overarching goal of this pilot study was to develop a screening method for the early detection of CRC and advanced adenoma.

## Results

### Patient Recruitment and Demographics

The study was conducted following a double-blinded design with recruitment at NorthShore University Health System, University of Chicago, and Indiana University. Of the 135 patients in our control group, 13 patients had hyperplastic polyps and 122 patients had other non-significant findings, and our case group consisted of 13 patients with diminutive adenoma (DA), 15 patients with nondiminutive adenoma (NDA), 74 patients with advanced adenoma (AA), 9 patients with hereditary non-polyposis CRC (HNPCC), and 10 patients with CRC. Patient demographic information collected included age, gender, smoking and drinking history. To evaluate potential confounding factors, we performed analysis of covariance (ANCOVA) on both control and case groups (defined as NDA, AA, Cancer) with the results shown in Fig. 1. The percentage of females was comparable between control (49%) and case (48%) groups. The proportion of smokers was slightly higher in the cancer population, whereas the percentage of drinkers was slightly higher in the control population.

**Figure 1.**
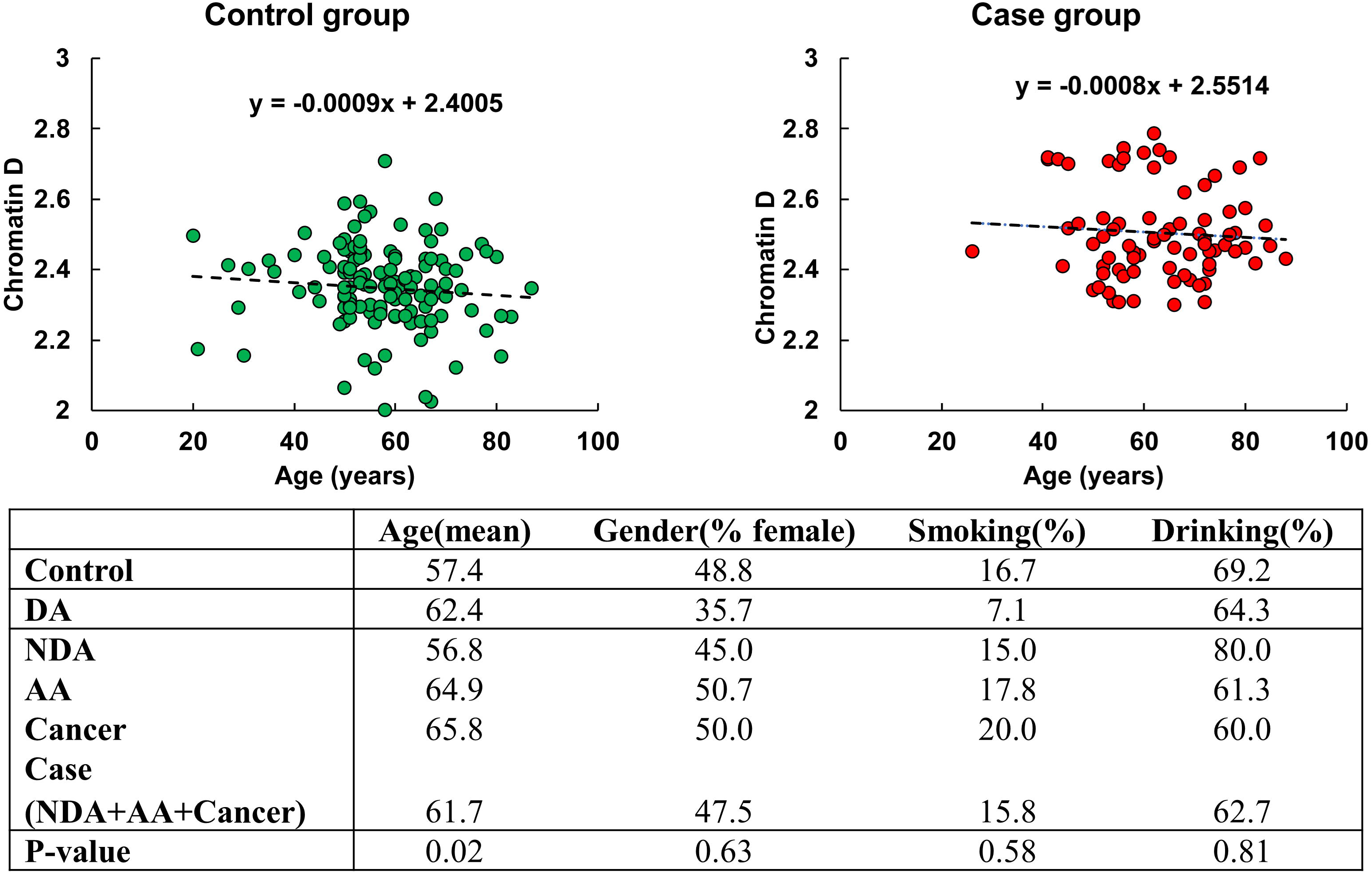
Demographic factors across different diagnostic endpoints. Case consists of nondiminutive adenoma (NDA), advanced adenoma (AA), and cancer groups.

ANCOVA analysis did not show any significant relationship between gender, smoking history, or drinking history and chromatin packing scaling D. Age was significantly higher in the case group with a mean of 62 years old compared to the control population with a mean of 57 years old and showed a small negative correlation (linear regression coefficient = −0.008) with D using the linear regression model. This suggests a minimal influence of age on rectal D, as a 10-year difference in age contributes to less than 7.2% of the variation in average D between the control and case populations, and, importantly, despite being on average slightly older, the cases had an elevated D compared to controls.

### csPWS is sensitive to chromatin domain alterations associated with field carcinogenesis

We investigated the influence of field carcinogenesis on chromatin structure by analyzing the packing scaling behavior of PDs of colonocytes brushed from different locations within the colorectal track. Our study focused on comparing samples obtained from the tumor site, normal appearing colonocytes brushed at locations 4 cm away from the tumor, and rectal colonocytes from patients with tumors. We observed a significant increase in D within nuclear chromatin domains in samples obtained from the tumor site, locations 4 cm away from the tumor, and the rectum compared to rectal colonocytes obtained from healthy controls (shown in Fig. 2a). However, no statistically significant differences were observed in D among the three tumor-associated locations (tumor, 4 cm away, and rectum). This suggests that our biomarker derived from rectal mucosa carries a distinct signature of field carcinogenesis which is robust throughout the colorectal tract.

**Figure 2.**
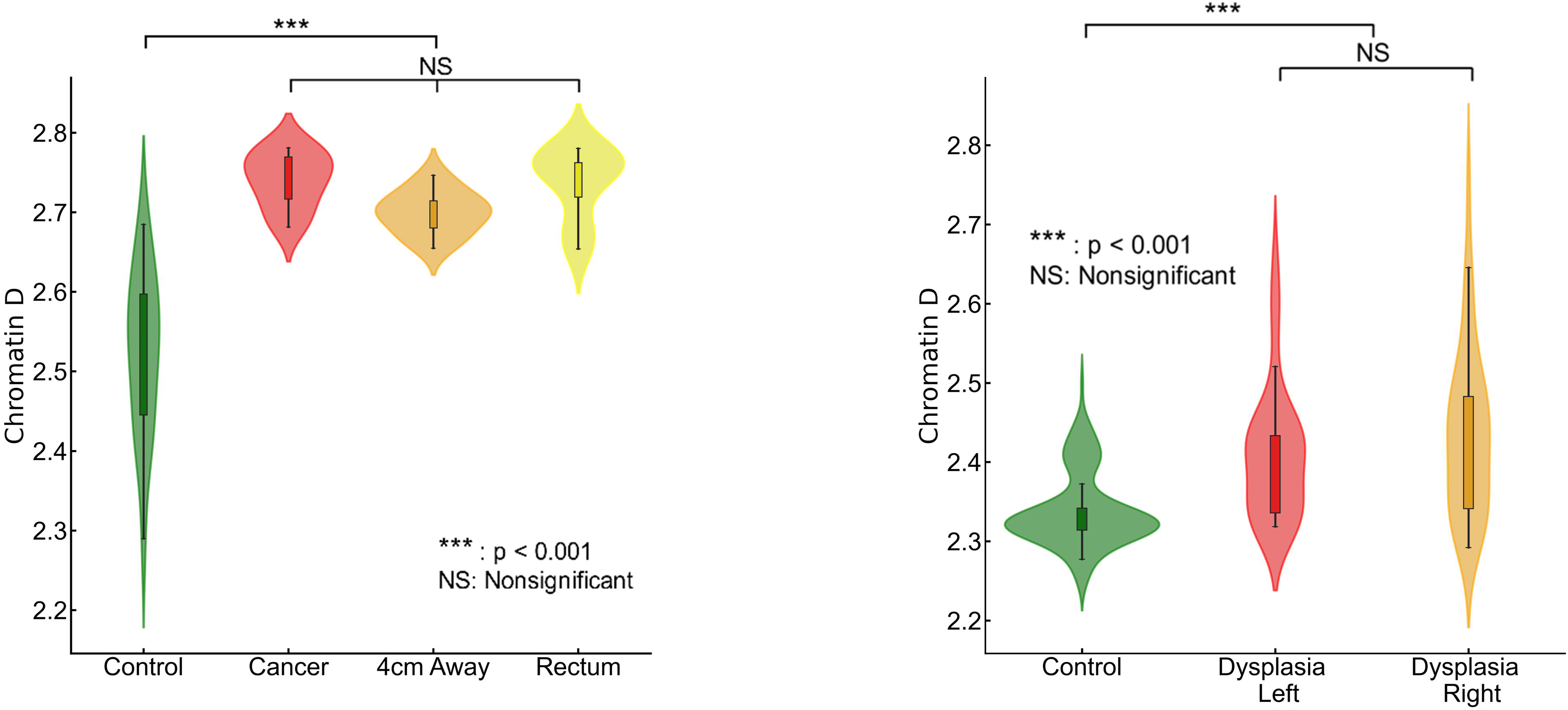
Packing scaling D is sensitive to field carcinogenesis. **(a)** Chromatin packing scaling D in cells brushed from tumor site, healthy-appearing tissue located 4 cm away from tumor and from rectum showed significantly increase (p=1.5×10^-6^,6.9×10^-5^, 3.6×10^-7^ respectively) compared to control patients but no significant difference among the three locations. **(b)** Rectal D is increased in patients with dysplasia regardless of anatomic location, right-sided (p=0.017) and left-sided adenoma (p=0.002) compared to control.

We assessed the effectiveness of rectal D as a potential biomarker for field carcinogenesis. In a separate dataset (135 controls and 74 cases), we observed that both left-sided and right-sided adenomas displayed a statistically significant increase in rectal D compared to the control group (Fig. 2b). This finding underscores D as a robust biomarker that is not limited by the location of an adenoma within the colon and rectum. Overall, our findings validate that chromatin structural changes measured by packing scaling D are indicative of field carcinogenesis in early-stage CRC patients regardless of the exact location of an adenoma.

### Chromatin PD alterations correlate with CRC risk

Prior studies on etiological field carcinogenesis highlighted the role of a preconditioned “field” in fostering transcriptomic, genomic, and epigenetic alterations that may lead to a neoplasm in the affected region. Therefore, the entire “field of injury” may bear the molecular biomarker of carcinogenesis irrespective of proximity to a tumor. Our objective was to detect nanoscale chromatin structural changes and alterations in PDs of rectal histologically normal appearing colonocytes that may serve as biomarkers of carcinogenesis and are detectable by csPWS. Our findings, as illustrated in Fig. 3, reveal a clear correlation between an increase in packing scaling D and colonoscopic findings. The rectal D measured from patients with abnormal colonoscopy findings (adenoma size > 5mm, HNPCC, or cancer) was significantly increased compared to rectal D measure from patients with a normal colonoscopy result. Specifically, we observed a non-significant increase in rectal D for smaller adenomas, such as diminutive adenoma (polyp size < 5 mm, n = 13). However, a significant increase in D was noted in patients harboring nondiminutive/nonadvanced adenomas (5-9 mm polyps, n = 15) and advanced adenomas (polyp size ≥ 10 mm, high-grade dysplasia or >25% villous features, n = 74). Moreover, rectal D was further elevated in patients with hereditary nonpolyposis colorectal cancer (HNPCC, n=9), characterized by a lifetime risk of CRC ranging from 60% to 80%, as well as in patients with colorectal cancer (n=10). Rectal D mirrored current and past colonoscopic findings and progressively increased from the low-risk CRC group to the high-risk CRC group: control < control with high-risk history < no-risk history with advanced adenoma < low-risk history with advanced adenoma < high-risk history with advanced adenoma (Fig. 4a). These results indicate that an increase in the putative biomarker has a robust correlation with the severity of precancerous lesions and CRC elsewhere in the colon.

**Figure 3.**
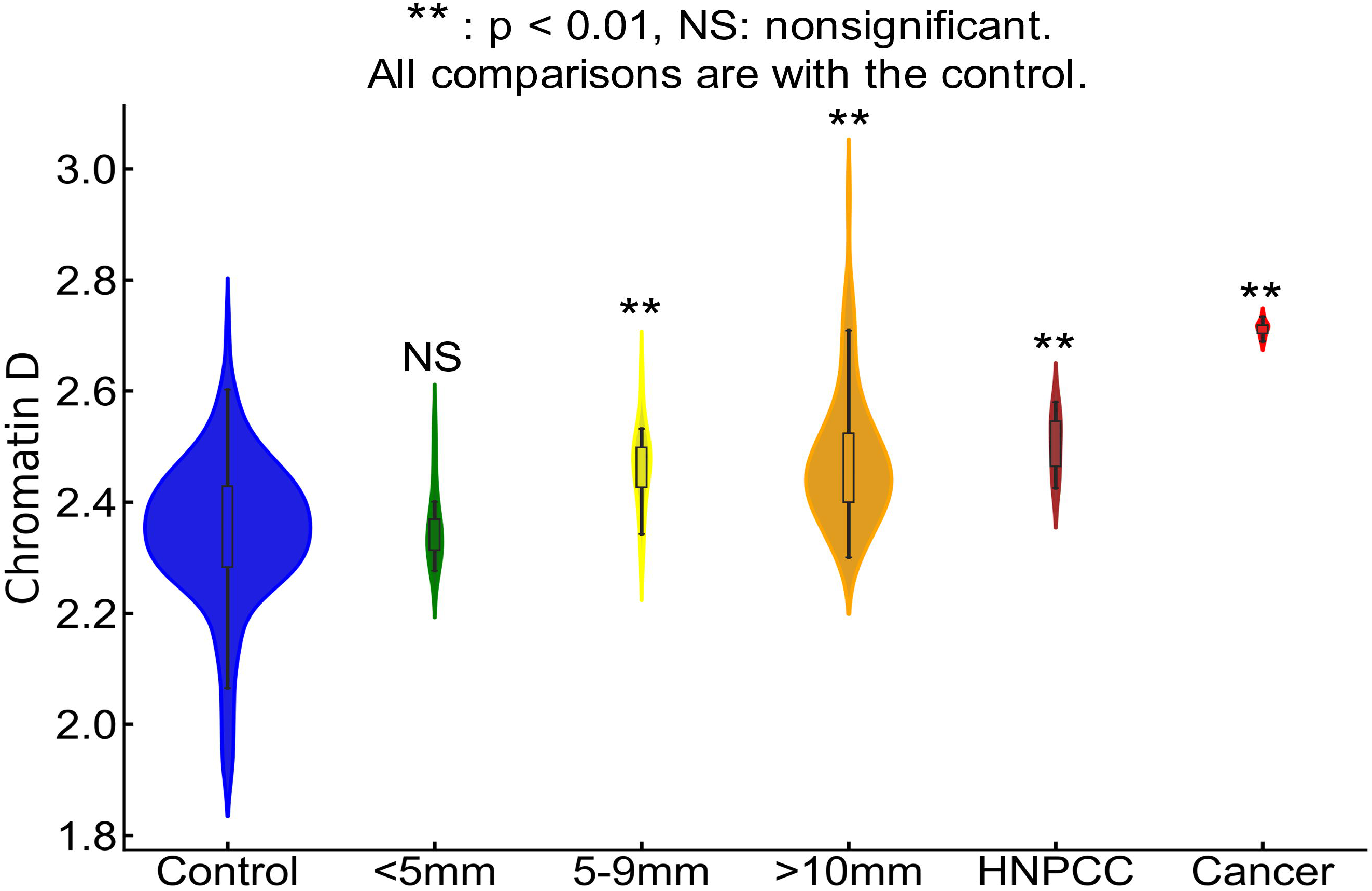
Rectal chromatin domain changes are sensitive to progression of CRC. Rectal D is increased progressively from control < diminutive adenoma (<5mm) < nondiminutive adenoma (5-9mm) < advanced adenoma (>10mm) < HNPCC < Cancer.

**Figure 4.**
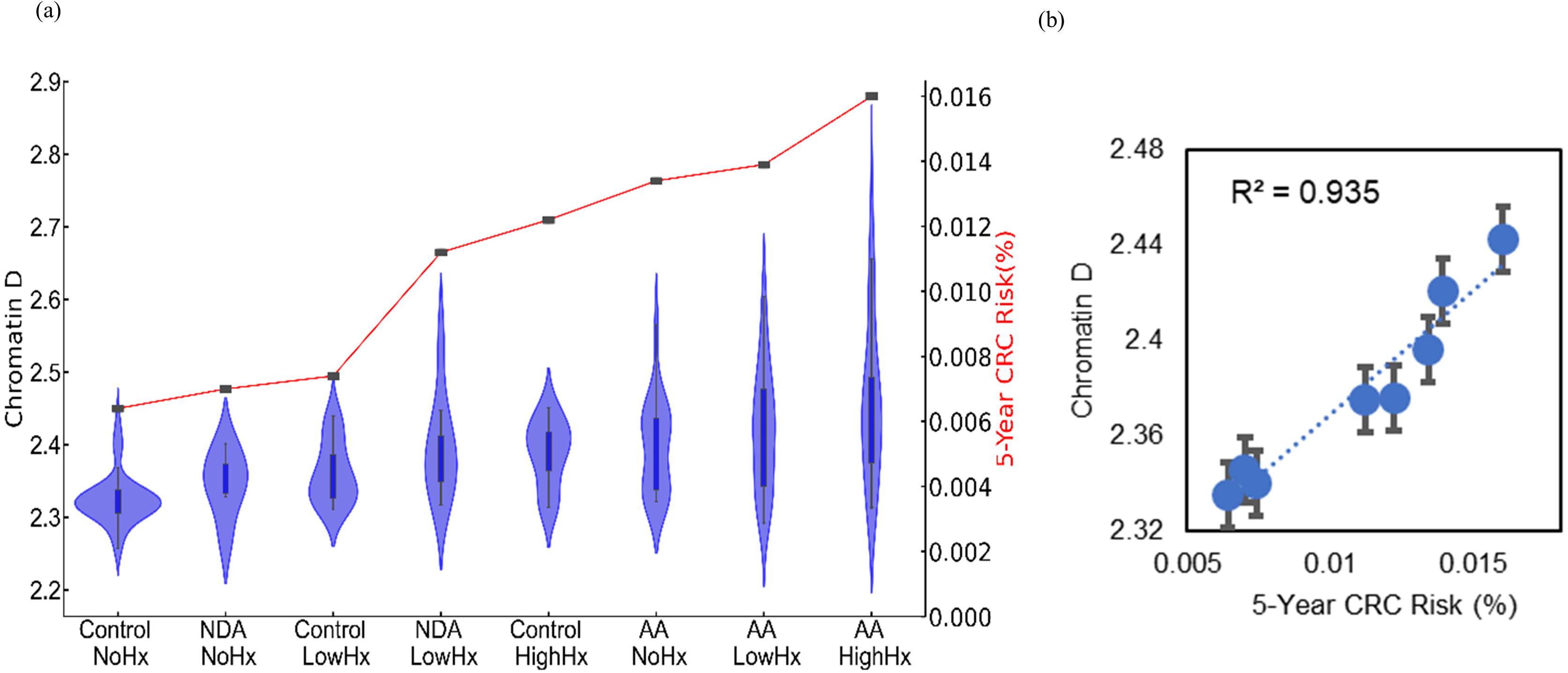
(a) Chromatin structural changes estimated by csPWS Rectal D correlated with colonic risk history. **(b)** 5-year cumulative CRC risk model and packing scaling D regression analysis, r^2^ = 0.94.

To assess the relationship between the dysregulation of chromatin PD in field carcinogenesis and the risk of CRC, we developed a five-year CRC risk model reflecting different stages of tumorigenesis (Fig. 4a). Rectal D effectively mirrored the risk of CRC progression. A statistically significant increase in rectal D was observed in high-risk advanced adenoma (effect size = 0.83), low-risk advanced adenoma (effect size = 0.79), and high-risk control populations (effect size=0.75) compared to low-risk and control populations without a history of CRC (Fig. 4a). Furthermore, regression analysis (Fig. 4b) revealed a positive correlation between packing scaling D and five-year CRC risk, demonstrating a strong correlation (r^2^ = 0.95). These findings demonstrate a robust and significant correlation between the dysregulation of chromatin in rectal colonocytes and the risk of CRC progression. The effectiveness of leveraging average packing scaling D in the detection of dysregulation of chromatin PD that may eventually contribute to the development of CRC provides the rationale for its use as a biomarker for CRC screening.

### csPWS-measured rectal D is sensitive to advanced adenomas throughout the colorectal tract

We obtained rectal brushings from the histologically normal mucosa of patients prior to colonoscopy (135 control, 74 advanced adenomas). The dataset was 50/50 split for prediction rule development and prospective testing. In the testing set, 0.85 sensitivity and 0.85 specificity with AUC = 0.85 were observed for control patents vs those with advanced adenomas located elsewhere in the colon. One crucial aspect that many early screening tests for CRC must consider is whether sensitivity is maintained for small lesions. We evaluated the proportion of advanced adenoma patients with different polyp sizes to test whether rectal D is limited by tumor load or lesion size (Fig. 5). The majority of the advanced adenoma lesions (78.4%) were under 1.5cm while only 5.4% were over 3 cm in size.

**Figure 5.**
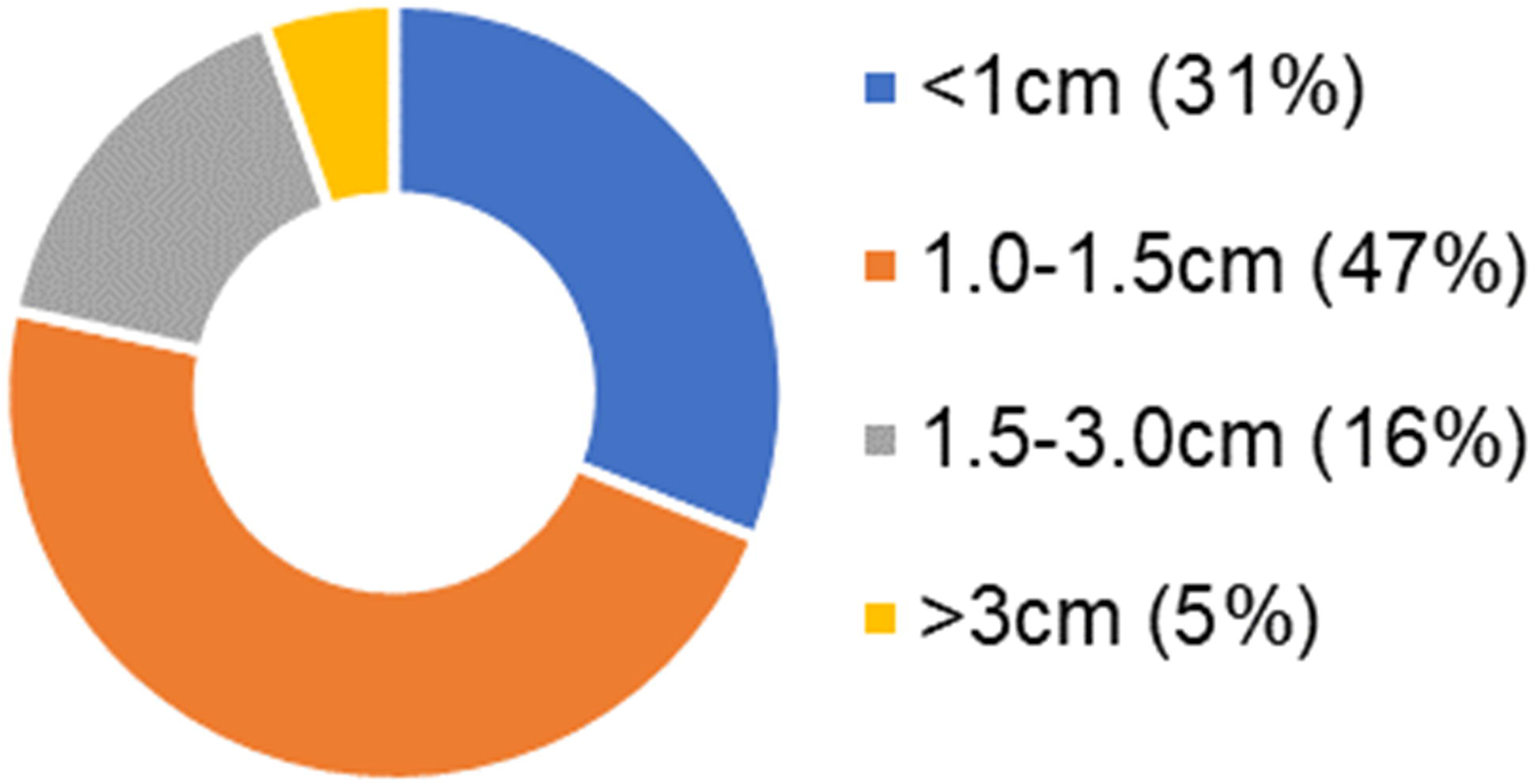
Proportion of patients with different lesion size in advanced adenoma group

### AI-enhanced csPWS analysis of chromatin alterations in rectal colonocytes provides improved diagnostic performance for detection of advanced adenomas

The complex link between physical chromatin organization and genetic/epigenetic alterations in early cancer development includes the association between gene expression and packing scaling D. Transcription involves a series of chemical reactions that are modulated through the balance between reaction rate constant and molecular accessibility of transcriptional reactants (RNA polymerase, transcriptional factors, etc.) and are affected by the local chromatin environment within packing domains. Leveraging recent advances in AI, specifically using convolutional neural networks, we utilized transfer learning paired with dimensionality reduction with an autoencoder network to better capture this complexity. The representative features were used on a random forest classifier, and the performance of the trained model was evaluated using the repeated stratified cross-validation sets (75/25 training/testing split). Optimal sensitivity and specificity values were selected based on the cut-point on the AUC curve that maximizes the number of correct classifications within each cross fold. Enhanced diagnostic performance in differentiating control and case populations was observed with AUC of 0.90 (+/−0.06), 0.88 (+/−0.08) sensitivity, and 0.85 (+/−0.09) specificity (Fig. 6). We also evaluated the diagnostic performance of the AI model for different endpoints (Table 1). Identical network structure was applied to different datasets with different subgroups categorized into controls and cases. These results show that AUC from our cross-validated model maintains robust diagnostic performance across different stages of CRC progression.

**Figure 6.**
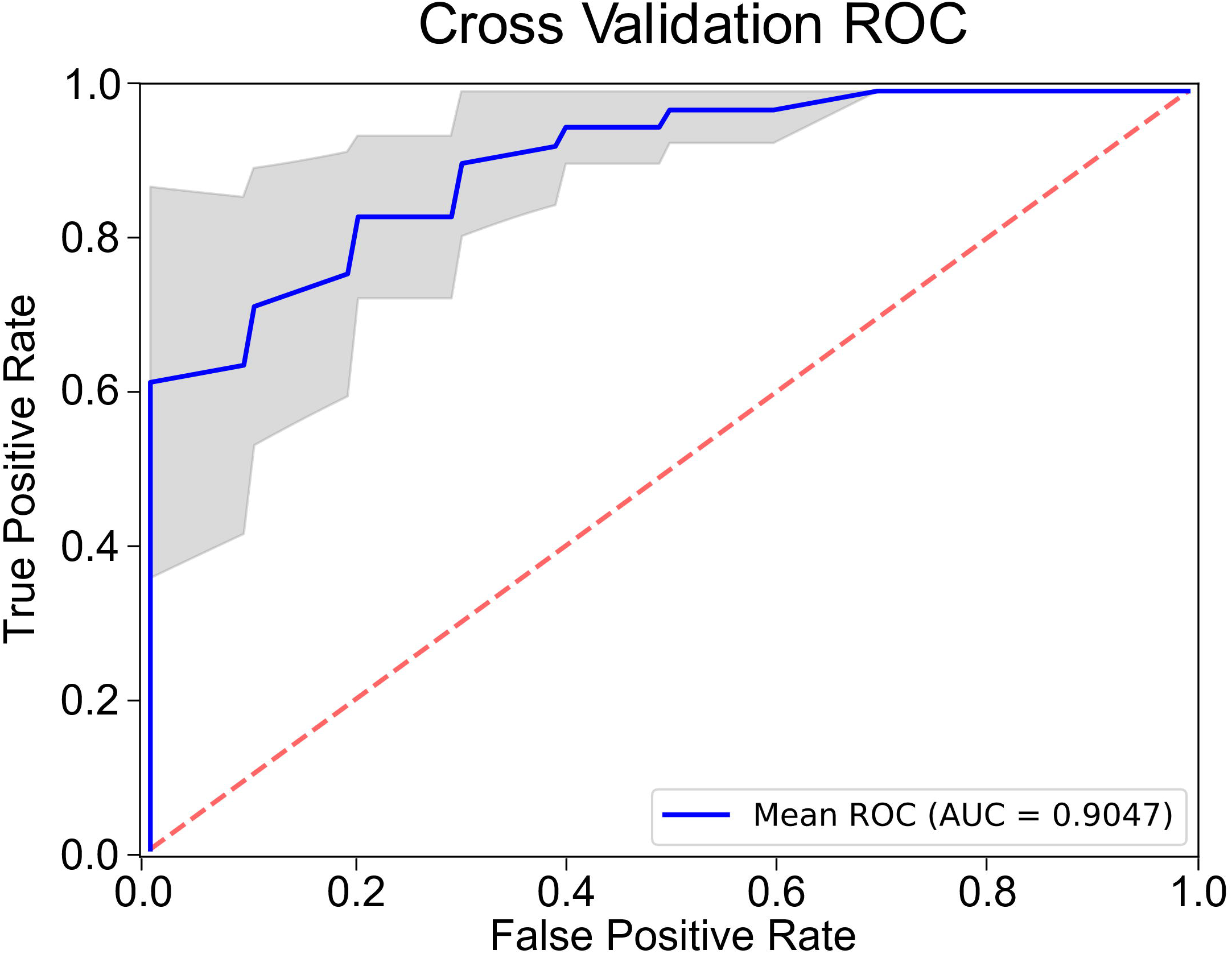
Diagnostic performance of AI-enhanced csPWS analysis of chromatin domain alterations in advanced adenoma. Blue AUC curve: mean for all cross-folds. Gray area shows 95% CI.

**Table 1.**
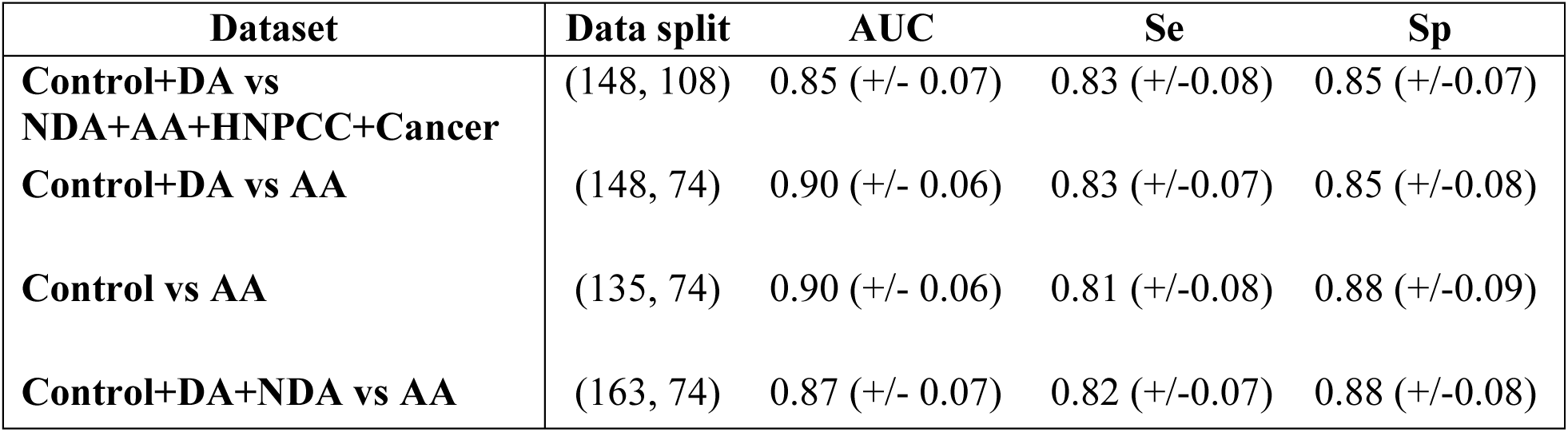
Diagnostic performance of AI model at different endpoints.

An important question is whether AI-enhanced csPWS is robust for identifying patients harboring advanced adenomas regardless of size. Implementing the previously discussed AI-enhanced analysis on subgroups of advanced adenoma based on lesion size (< 1cm, 1-1.5 cm, and >1.5cm), a comparable classification performance was achieved for lesions of different sizes. With a fixed specificity of 0.88, the sensitivity of successfully identifying advanced adenoma ranged from 0.81 to 0.83 (Table 2). Our AI-enhanced csPWS thus demonstrated the ability of our proposed biomarker to detect small lesions by leveraging the characteristics of field carcinogenesis, enabling early detection of CRC and advanced adenoma.

**Table 2.** Diagnostic performance of AI model in subgroups of advanced adenoma based on lesion size.

| Category | N (%) in subgroups of AA | Sensitivity |
| --- | --- | --- |
| < 1 cm | 23 (31.1) | 0.83 |
| 1-1.5 cm | 35 (47.3) | 0.83 |
| > 1.5 cm | 16 (21.6) | 0.81 |

## Discussion

Most CRCs arise from adenomatous colon polyps that progress into advanced adenomas and then to carcinomas. Screening from 45 years of age is recommended for average-risk patients with the goal of improved disease prognosis by identifying an early stage of CRC that is more treatable, resulting in a reduced mortality rate[10]. Among the multiple tests available for early screening of CRC, each modality has its own limitations: stool-based tests have low sensitivity, CT colonography involves radiation, and endoscopy is costly and requires bowel preparation and sedation [4, 5]. The novel liquid biopsy tests that are being developed by companies including Grail, Freenome, Guardant, Delfi, and Thrive show excellent results in detection of CRC but suboptimal performance in identifying advanced adenoma[17–25]. This suboptimal performance in detecting early-stage cancers and precancerous significant lesions can be attributed to the biological nature of the biomarker source. Circulating cell-free DNA as a biomarker poses the limitation of its small fraction within the peripheral blood, with the majority of DNA originating from hematopoietic cells[26, 29]. Early-stage cancer development is likely associated with a smaller oncogenic load that will result in a smaller amount of biomarker released into the blood stream. It is thus likely that sensitivity to early oncogenesis drops due to fewer genetic/epigenetic alterations being accumulated, resulting in less heterogeneity within the clonal expansion process that these tests utilize.

To overcome the loss of sensitivity for smaller lesions that plague many current tests, we explored an alternate biomarker source and type. Our results suggest that field carcinogenesis is a promising biomarker source for early CRC detection. Field carcinogenesis implies that extensive epigenetic alterations preceding dysplastic changes are not limited to the tumor site alone but encompass the entire “field of injury”, regardless of the tumor load [50]. Thus, by combining the biomarker source of field carcinogenesis and biomarker of chromatin structural changes, we expect a highly sensitive and reliable modality for identifying early CRC development.

Previous studies show both experimentally and computationally that chromatin packing scaling D, the size of chromatin domains, and chromatin density all affect the local macromolecular crowding and may play a crucial role in the regulatory mechanisms expressing phenotypic plasticity[40, 43]. It has been proposed that an increase in phenotypic plasticity of gene expression can be linked to carcinogenesis with potential mechanisms involving neoplastic cells’ increased chance of survival in response to external stressors by modulation of transcriptional malleability and intercellular transcriptional heterogeneity. Detection of chromatin conformation biomarkers can be achieved using csPWS, a high-throughput optical nanosensing technology that enables nanoscale detection of changes in chromatin domain conformation. Leveraging the clinical protocol that was developed for cell acquisition, storage, and shipment, a reliable measurement of physical characteristics of the chromatin structure can be performed on rectal colonocytes obtained from rectal brushings. With a length scale sensitivity of 23-334 nm, csPWS is optimized to sense PDs (average size of ~200 kbp), in which chromatin packing behavior can be characterized by packing scaling D.

Our findings demonstrate utilization of field carcinogenesis in CRC as a powerful tool for early colorectal cancer detection. We showed that rectal D measurements using csPWS are sensitive to field carcinogenetic and can be leveraged to differentiate healthy patients from those who harbor adenomatous lesions within the entire colon. Dysregulation of chromatin PD in colonocytes obtained from normal-appearing rectal tissue in patients with CRC, as well as those located 4 cm away from the tumor showed an increase in D compared to colonocytes from control patients. Our data show that rectal D is increased in patients harboring adenomas regardless of their location, at distal or proximal colon tract. These results suggest that chromatin biomarkers of field carcinogenesis can be obtained from rectal colonocytes. We confirmed the relationship between rectal D and the risk of progression to CRC via development of a risk stratification model based on colonoscopy findings. We developed a model of 5-year risk of progression to CRC based on colonoscopic findings and found a robust correlation between the dysregulation of chromatin in rectal colonocytes and the risk of progression. These results indicate that chromatin PD changes within the nucleus of rectal colonocytes mirror changes throughout the colon, demonstrating the potential of our proposed marker for early CRC screening with easy accessibility via rectal colonocyte brushings.

Our initial univariate analysis of using the nuclear average of packing scaling D of rectal colonocytes as a sole biomarker showed the ability to differentiate patients harboring advanced adenomas from control subjects with AUC=0.85. However, the average rectal D of chromatin packing domains may not fully capture the complexity of the interplay between chromatin conformation and regulation of gene expression. Domain size, chromatin volume concentration, domain volume fraction, histone marks, interdomain structure, and other properties of 3D chromatin structure have been shown to modulate the PD regulation of transcriptional plasticity. Consequently, we utilized an AI-based feature engineering approach to better capture the key information that chromatin structural changes may present[42]. Our AI-based model leverages the power of deep learning algorithms, specifically through transfer learning pre-trained on a large dataset from ImageNet. The transfer learning network enables the extraction of features with information that may be difficult to attain through different analytical approaches. Our network utilizes dimensionality reduction using an autoencoder to optimize the features more representative of our data. The resultant features were then passed onto our binary classification model for differentiating healthy from those with advanced adenoma. Our model’s robustness was validated using repeated stratified 4-fold cross-validation. The diagnostic performance was evaluated with AUC, sensitivity, and specificity metrics with excellent results of AUC = 0.90(+/−0.06), sensitivity = 0.88(+/−0.08), and specificity = 0.85(+/−0.09) for advanced adenoma. We should note that the sensitivity and specificity were selected based on the optimum point on the AUC curve within each cross fold. We would like to emphasize that a majority of the adenomas that were measured in our study were small in size (< 1.5cm), adding immense clinical value in the early prediction of CRC. Implementation of our model to the advanced adenoma subgroups based on lesion size showed comparable results with the accuracy of correctly identifying as harboring advanced adenoma from 81% to 83%. As our model is not dependent on tumor load, early changes manifested in chromatin nanostructures under prolonged field injury may serve as a new opportunity for a sensitive early screening tool.

We have shown that the clinical protocol of rectal colonocyte acquisition and csPWS imaging, further aided by AI-based feature engineering, can provide a sensitive modality for the detection of advanced adenoma. Our study was constrained by certain limitations, however. The study recruited a limited number of patients; therefore, it cannot provide a definitive evaluation of our approach’s performance. All subjects were undergoing screening or surveillance colonoscopy; however, the ratio of cases compared to healthy control in our study are notably higher than the disease prevalence among the screening population. Future risk prediction modeling can be extended from the current study once our model is shown to be robust across different demographic populations with larger-scale recruitment. The possible impact of other confounding factors such as age, dietary and lifestyle habits should be further evaluated, and any effect of potential small debris or mucus on the csPWS signal may also be investigated.

## Material and Methods

### Patient Recruitment

All studies performed and samples collected were under the approval of the Institutional Review Board at NorthShore University Health System, the University of Chicago, and Indiana University. All methods were performed in accordance with the relevant guidelines and regulations and written informed consent was obtained from all participants undergoing screening or surveillance colonoscopy. The exclusion criteria for recruitment included incomplete colonoscopy due to failure to visualize the cecum or patients with coagulopathy, past medical history of pelvic radiation, or systemic chemotherapy. Patient demographic information including age, sex, smoking and drinking history were gathered. The diagnostic criteria for each and all subjects were made by a board accredited GI specialist and pathologist based on colonoscopy and pathology reports.

### Sample collection and shipment

All sample acquisitions were adherent to the following minimally invasive protocol: colonoscopy to cecum was performed with standard techniques using Olympus 160 or 180 series or Fujinon colonoscopes. A sterile cytology brush (Cytobrush, CooperSurgical, Inc., Trumbull, CT, USA) was passed through the endoscope after insertion into the rectum, and gentle pressure with rotation of bristle was applied to the rectum. A single cytology brush was used for each patient, and the tip of the brush was clipped and immediately immersed in 1.5 mL vile tube filled with 750 mL of 25% ethanol. The samples were packaged and shipped to Northwestern University on the same day. Temperature was maintained below 10°C with polar pack refrigerant gel (SONOCO Thermosafe, Arlington Heights, IL, USA), and packaging was adherent to guidelines provided by the Department of Transportation with a primary and secondary container with absorbent material.

### Sample deposition and preparation

All sample deposition and preparation were performed by an investigator blinded to patient information: Within 24 hours of sample acquisition, the brush was smeared onto two microscope glass slides (Fisher Scientific, Hampton, NH, USA), which were then fixed in 95% ethanol for 30 minutes. The slides were examined under a bright field microscope to find cells deposited onto the cytology slide consisting of different types of cells including epithelial cells, red blood cells, and inflammatory cells. All measurements were taken from columnar epithelial cells as identified by standardized hematoxylin and cytostain staining protocol. Samples with sufficient columnar epithelium free of crest, fold, cell debris, and mucus were only included in the study and imaged with csPWS. Based upon power analysis performed with confidence interval (CI) on average D restricted to be less than 5% of the difference between control and case populations, the minimum number of cells collected was set to >30 cells per patient.

### csPWS Instrumentation and Imaging

The csPWS instrument was built on a commercial microscope (Nikon Instruments, Melville, NY, USA) with modifications to include a Xenon lamp (Oriel Instruments, Stratford, Connecticut, USA). The spatially incoherent white light was focused onto the sample and a back-scattered image is projected through a liquid crystal tunable filter (Cri, Woburn, MA, USA) with a spectral resolution of 7 nm and further onto a CCD camera (Princeton Instruments, Trenton, NJ, USA). Monochromatic spectrally resolved images of wavelengths within 500-700 nm (at 2 nm increments) are acquired with the resulting data stored in an image cube (x, y, λ) and normalized by the reference wave acquired at a blank region on the slide. We used a moderately small numerical aperture (NA) of light incidence of 0.6, and light collection NA of 0.8 for csPWS to produce a uniform intensity across the sample plane. csPWS achieves sensitive but non-resolvable sub-diffraction length scale of chromatin in the range of 23 – 334 nm. Within the nucleus, the refractive index (RI) is proportional to the local macromolecular density *ρ*(r) mainly consisting of protein, DNA, RNA, and others. The refractional increment is constant and mainly contributed by chromatin and nearly independent of the chemical constituents.

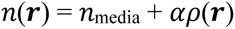

The readout of PWS microscopy is the image of a cell that captures and quantifies spatial fluctuations in macromolecular density via evaluating the standard deviation of the interference spectra (Σ) between the spectrum of the reference wave and the scattering caused by the spatial variations of *ρ*(***r***) across different wavelengths. The value of Σ is proportional to the Fourier transform of the autocorrelation function (ACF) of *ρ*(***r***), which is integrated over the Fourier transform of the coherence volume. Coherence volume was defined by the spatial coherence in the transverse direction (458×458 nm^2^) and the depth of field in axial direction (~3 µm). Consequently, the range of length scale sensitivity of the spectral interference signal and Σ depend on the illumination and collection geometry of the instrument, in particular their numerical apertures and the spectral bandwidth. We chose these instrument parameters to maximize the sensitivity of the interference signal to the length scales relevant to chromatin conformation within packing domains. As the fundamental unit of PDs is the 5-20 nm chromatin chain, the average domain diameter is 160 nm, and larger domains approach 400 nm in diameter, the instrument parameters were chosen such that the interference signal is predominantly sensitive to chromatin density variations at length scales from approximately 23 to 334 nm. For each intranuclear location (x,y), Σ(x,y) was used to calculate chromatin packing density scaling D(x,y) using the previously reported algorithm [49]. In particular, we employed an analytical framework that integrates finite difference time domain simulation and experimental results to determine the packing scaling parameter D for each pixel within a 458 nm by 458 nm area based on Σ [35]. Chromatin is the strongest contributor to the csPWS signal within the nucleus, as most other mobile macromolecules are outside the length-scale sensitivity of csPWS. In this analytical framework, the packing scaling parameter D was calculated by fitting the mass-density autocorrelation function (ACF) obtained from Σ measurements in PWS to the ACFs obtained from ground truth measurements of chromatin structure in lung adenocarcinoma A549 cells and differentiated BJ fibroblasts using chromatin transmission electron microscopy (ChromTEM) images [49]. In short summary, the Σ(x,y) is proportional to the spatial ACF of the mass density distribution, B(r), convolved with a smoothing function S(r), which is characterized by the optical system setup and the source spectrum. We should note that S(r) thus depends on various factors including numerical aperture of the microscope, sample characteristics of the cell such as density of chromatin and macromolecular crowding, chromatin volume concentration, genomic lengths, and sample-glass interface characteristics such as forward and reverse Fresnel reflection and transmission coefficients and refractive index of media and nucleus. A model parameter D_b_ that describes the shape of B(r) can be obtained for each given Σ within each coherence volume, which enable us to calculate the packing scaling D using the following relationship.

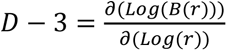

The estimation of packing scaling D took into account the influence of chromatin volume concentration ϕ and genomic size Nf of packing domains. By considering these factors, the framework allowed for a more accurate determination of the packing scaling behavior within the chromatin structure.

### Evaluation of average packing scaling D

We investigated the influence of field carcinogenesis on the packing scaling behavior of chromatin PDs within the nucleus of rectal mucosa. We compared a total of 201 patients, comprising three groups: controls (n=136), patients with right-sided adenoma (n=27), and patients with left-sided adenoma (n=38). Tissue samples were collected from various distances relative to the tumor tissue, including samples obtained directly from the tumor as well as tissues located 4 cm away from the tumor and rectum. These samples were compared to tissues collected from a healthy control population. Using PWS microscopy, we quantified the average packing scaling parameter D in the nucleus of rectal mucosa for each sample group. By comparing these values across different distances from the tumor and with the control group, we aimed to assess the impact of field carcinogenesis on the chromatin PDs within the rectal mucosa.

### CRC 5-year risk model

In addition to our investigation of chromatin PDs, we also developed a CRC risk model that aims to estimate the cumulative 5-year risk of developing CRC for different populations based on their baseline colonoscopy and follow up surveillance colonoscopy. The risk model is built upon published data from a consensus update provided by the US Military-Society Task Force and a study by Pinsky et. al. on surveillance. To construct the risk model, we divided the study population within our dataset into three categories: no history, low-risk history, and high-risk history based on past surveillance colonoscopy findings. By considering both baseline colonoscopy and current colonic health, we developed a cumulative 5-year risk model by incorporating the following factors: annual risk of nonsignificant finding or diminutive adenoma progression into advanced adenoma, the annual risk of CRC progression from advanced adenoma, and the risk of developing metachronous CRC into the model.

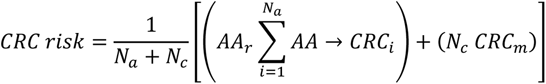

where Na is number of patients with no history or history of adenoma, Nc is number of patients with history of cancer, AAr is the cumulative risk of developing future advanced adenoma, AA→CRC is the risk of AA to CRC, and CRCm is the cumulative risk of developing metachronous CRC. It should be noted that we follow the results from US Military-Society Task Force that the risk progression in CRC depends both on sex and age, therefore calculating individual annual risk progressions in different sub-categories (male vs female, age below and above 80 years old). The annual risk progression from AA to CRC is converted into cumulative risk using the following formula.

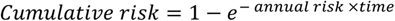

By incorporating these key factors, our risk model provided a tool for a comprehensive evaluation of the impact of packing scaling D and chromatin structural changes during the progression and development of CRC, including early stages such as adenoma. We leverage this 5-year cumulative risk model as a reference to evaluate whether rectal D is sensitive to field carcinogenesis, not restricted to the active level of dysplasia but also to the past colonoscopy results representative of field injury on the system.

### AI analysis of packing scaling D

AI was employed to assess the potential of packing scaling D as a putative biomarker for early detection of CRC and advanced adenoma. A deep learning approach was leveraged to capture the complex relationship between D, a physical descriptor of chromatin organization, and oncogenic transformation.

Our AI-driven approach consisted of four steps: nucleus segmentation, preprocessing, feature learning, and classification (shown in Fig. 7). Nucleus segmentation was conducted by a trained investigator using custom software with graphic user interface, while remaining blinded to the patient information. The segmented D images on nuclei were resized and subjected to min-max normalization during the pre-processing step.

**Figure 7.**
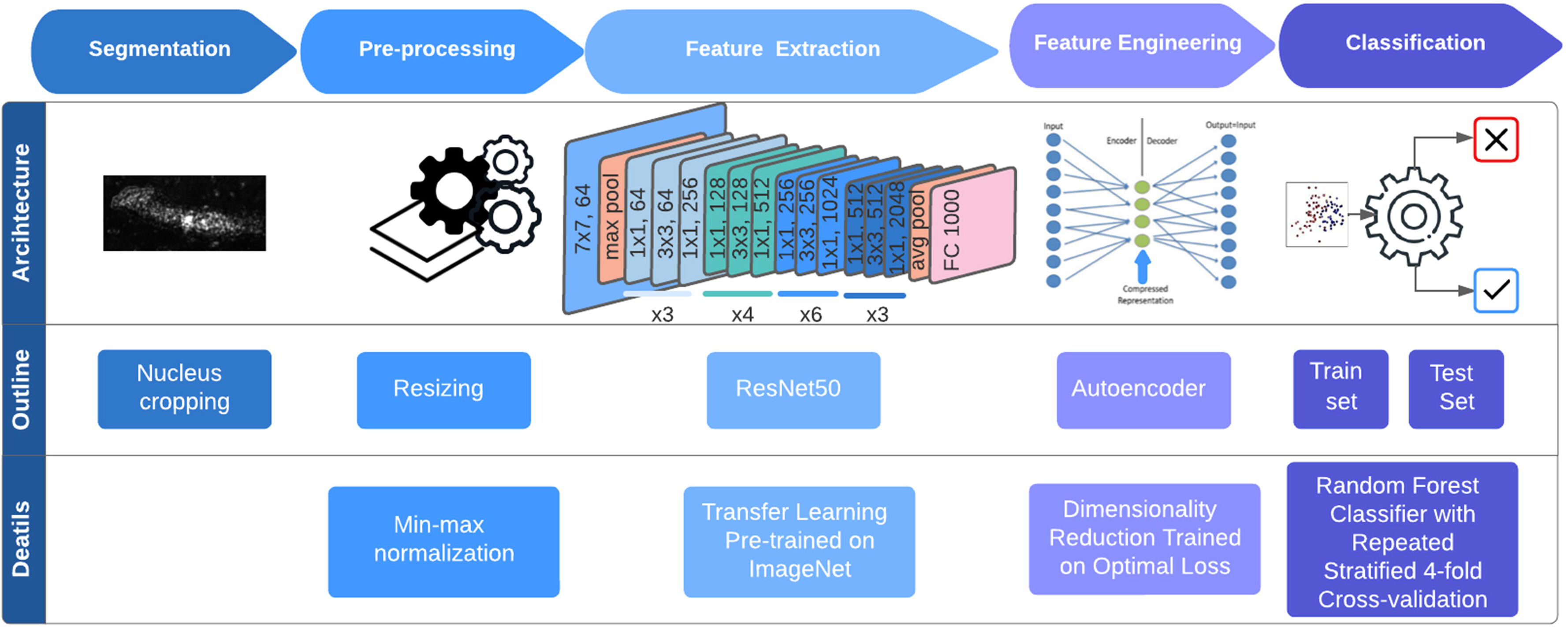
Workflow and architecture of the AI driven feature engineering model

For feature learning, we employed a transfer learning approach with ResNet50, a convolutional neural network (CNN) pretrained on ImageNet database. Features were extracted from the final convolutional layer of the CNN architecture. To enhance data representation and computational efficiency, an autoencoder network was implemented. The autoencoder was trained to minimize the optimal loss, and the encoder output served as representative feature.

In the classification step, a binary classification using a parameter-tuned random forest classifier was implemented on the training set to distinguish the healthy control population from the case population with advanced adenoma. The classifier model was fine-tuned through grid search, exploring multiple configurations, and selecting one with minimal error on our dataset. To robustly evaluate our performance on relatively small dataset, we employed a repeated stratified 4-fold cross-validation method with five iterations to compute our diagnostic performance on metrics including area under the curve (AUC), sensitivity, and specificity. Optimal sensitivity and specificity values were selected based on the cut-point on the AUC curve that maximizes the number of correct classifications within each cross fold. By repeatedly splitting the data into four folds and iteratively evaluating the results, we obtained reliable estimates of our diagnostic performance across different subsets of the dataset. This rigorous evaluation method enhances the generalizability and reliability of our findings.

### Code Availability

All computer codes used for the analyses in the study are available from the corresponding author on reasonable request.

## Data availability

The raw datasets generated and/or analyzed during the study are available from the corresponding author on reasonable request.

## Data Availability

All data produced in the present work are contained in the manuscript

## Acknowledgements

This study was supported by National Institutes of Health (NIH) grants U54CA268084, R01CA228272, R01CA225002, and NSF grant EFMA-1830961 with philanthropic support from Rob and Kristin Goldman, David Sachs and the Christina Carinato Charitable Foundation. AC wishes to thank Benjamin D Keane for his valuable assistance in manuscript preparation.

## Author contributions

V.B., H.S., and H.K.R. designed the project. A.C. and S.P. contributed equally to data analysis. A.C. and A.D. prepared the manuscript, and all authors contributed, providing feedback.

## Ethics declarations

Drs. Backman, Subramanian, and Roy are cofounders and/or shareholders in NanoCytomics LLC. All aspects of this study were done under the supervision of the Conflict of Interest Committee at Northwestern University.

## Reference

1. Sung, H., et al., Global Cancer Statistics 2020: GLOBOCAN Estimates of Incidence and Mortality Worldwide for 36 Cancers in 185 Countries. CA Cancer J Clin, 2021. 71(3): p. 209–249.

2. SEER. *Cancer Stat Facts: Colorectal Cancer*. 2020; Available from: https://seer.cancer.gov/statfacts/html/colorect.html.

3. Siegel, R.L., et al., Colorectal cancer statistics, 2023. CA Cancer J Clin, 2023. 73(3): p. 233–254.

4. Warren, J.L., et al., Adverse events after outpatient colonoscopy in the Medicare population. Ann Intern Med, 2009. 150(12): p. 849–57, W152.

5. Rabeneck, L., et al., Bleeding and perforation after outpatient colonoscopy and their risk factors in usual clinical practice. Gastroenterology, 2008. 135(6): p. 1899–1906, 1906 e1.

6. Ahnen, D.J., et al., The increasing incidence of young-onset colorectal cancer: a call to action. Mayo Clin Proc, 2014. 89(2): p. 216–24.

7. Eng, C. and H. Hochster, Early-Onset Colorectal Cancer: The Mystery Remains. J Natl Cancer Inst, 2021. 113(12): p. 1608–1610.

8. Robertson, D.J., et al., Recommendations on Fecal Immunochemical Testing to Screen for Colorectal Neoplasia: A Consensus Statement by the US Multi-Society Task Force on Colorectal Cancer. Gastroenterology, 2017. 152(5): p. 1217–1237 e3.

9. Lee, J.K., et al., Accuracy of fecal immunochemical tests for colorectal cancer: systematic review and meta-analysis. Ann Intern Med, 2014. 160(3): p. 171.

10. Lin, J.S., et al., Screening for Colorectal Cancer: Updated Evidence Report and Systematic Review for the US Preventive Services Task Force. JAMA, 2021. 325(19): p. 1978–1998.

11. Calistri, D., et al., Fecal multiple molecular tests to detect colorectal cancer in stool. Clin Gastroenterol Hepatol, 2003. 1(5): p. 377–83.

12. Lenhard, K., et al., Analysis of promoter methylation in stool: a novel method for the detection of colorectal cancer. Clin Gastroenterol Hepatol, 2005. 3(2): p. 142–9.

13. Hol, L., et al., Screening for colorectal cancer: randomised trial comparing guaiac-based and immunochemical faecal occult blood testing and flexible sigmoidoscopy. Gut, 2010. 59(1): p. 62–8.

14. Imperiale, T.F., D.F. Ransohoff, and S.H. Itzkowitz, Multitarget stool DNA testing for colorectal-cancer screening. N Engl J Med, 2014. 371(2): p. 187–8.

15. Diaz, L.A., Jr. and A. Bardelli, Liquid biopsies: genotyping circulating tumor DNA. J Clin Oncol, 2014. 32(6): p. 579–86.

16. Merker, J.D., et al., Circulating Tumor DNA Analysis in Patients With Cancer: American Society of Clinical Oncology and College of American Pathologists Joint Review. Arch Pathol Lab Med, 2018. 142(10): p. 1242–1253.

17. Liu, M.C., et al., Sensitive and specific multi-cancer detection and localization using methylation signatures in cell-free DNA. Ann Oncol, 2020. 31(6): p. 745–759.

18. Klein, E.A., et al., Clinical validation of a targeted methylation-based multi-cancer early detection test using an independent validation set. Ann Oncol, 2021. 32(9): p. 1167–1177.

19. Wan, N., et al., Machine learning enables detection of early-stage colorectal cancer by whole-genome sequencing of plasma cell-free DNA. BMC Cancer, 2019. 19(1): p. 832.

20. Ulz, P., et al., Inference of transcription factor binding from cell-free DNA enables tumor subtype prediction and early detection. Nat Commun, 2019. 10(1): p. 4666.

21. Cohen, J.D., et al., Detection and localization of surgically resectable cancers with a multi-analyte blood test. Science, 2018. 359(6378): p. 926–930.

22. Cristiano, S., et al., Genome-wide cell-free DNA fragmentation in patients with cancer. Nature, 2019. 570(7761): p. 385–389.

23. Chen, X., et al., Non-invasive early detection of cancer four years before conventional diagnosis using a blood test. Nat Commun, 2020. 11(1): p. 3475.

24. Kim, S.-T., et al., Abstract 916: Combined genomic and epigenomic assessment of cell-free circulating tumor DNA (ctDNA) improves assay sensitivity in early-stage colorectal cancer (CRC). Cancer Research, 2019. 79(13_Supplement): p. 916–916.

25. Raymond, V.M., et al., Evaluation of the ctDNA LUNAR-2 Test In an Average Patient Screening Episode (ECLIPSE). Journal of Clinical Oncology, 2021. 39(3_suppl): p. TPS142–TPS142.

26. Fiala, C. and E.P. Diamandis, Utility of circulating tumor DNA in cancer diagnostics with emphasis on early detection. BMC Med, 2018. 16(1): p. 166.

27. Campos-Carrillo, A., et al., Circulating tumor DNA as an early cancer detection tool. Pharmacol Ther, 2020. 207: p. 107458.

28. Bettegowda, C., et al., Detection of circulating tumor DNA in early- and late-stage human malignancies. Sci Transl Med, 2014. 6(224): p. 224ra24.

29. Junca, A., et al., Detection of Colorectal Cancer and Advanced Adenoma by Liquid Biopsy (Decalib Study): The ddPCR Challenge. Cancers (Basel), 2020. 12(6).

30. Putcha, G., et al., Blood-based detection of early-stage colorectal cancer using multiomics and machine learning. Journal of Clinical Oncology, 2020. 38(4_suppl): p. 66–66.

31. Lochhead, P., et al., Etiologic field effect: reappraisal of the field effect concept in cancer predisposition and progression. Mod Pathol, 2015. 28(1): p. 14–29.

32. Hawthorn, L., L. Lan, and W. Mojica, Evidence for field effect cancerization in colorectal cancer. Genomics, 2014. 103(2-3): p. 211–21.

33. Lewis, J.D., et al., Detection of Proximal Adenomatous Polyps With Screening Sigmoidoscopy: A Systematic Review and Meta-analysis of Screening Colonoscopy. Archives of Internal Medicine, 2003. 163(4): p. 413–420.

34. Atkin, W.S., et al., Once-only flexible sigmoidoscopy screening in prevention of colorectal cancer: a multicentre randomised controlled trial. Lancet, 2010. 375(9726): p. 1624–33.

35. Rebello, D., et al., Field carcinogenesis for risk stratification of colorectal cancer. Adv Cancer Res, 2021. 151: p. 305–344.

36. Suzuki, H., et al., Genome-wide profiling of chromatin signatures reveals epigenetic regulation of MicroRNA genes in colorectal cancer. Cancer Res, 2011. 71(17): p. 5646–58.

37. Bandres, E., et al., Epigenetic regulation of microRNA expression in colorectal cancer. Int J Cancer, 2009. 125(11): p. 2737–43.

38. Ng, J.M. and J. Yu, Promoter hypermethylation of tumour suppressor genes as potential biomarkers in colorectal cancer. Int J Mol Sci, 2015. 16(2): p. 2472–96.

39. Xu, J., et al., Super-resolution imaging reveals the evolution of higher-order chromatin folding in early carcinogenesis. Nat Commun, 2020. 11(1): p. 1899.

40. Li, Y., et al., Nanoscale chromatin imaging and analysis platform bridges 4D chromatin organization with molecular function. Sci Adv, 2021. 7(1).

41. Ou, H.D., et al., ChromEMT: Visualizing 3D chromatin structure and compaction in interphase and mitotic cells. Science, 2017. 357(6349).

42. Li, Y., et al., Analysis of three-dimensional chromatin packing domains by chromatin scanning transmission electron microscopy (ChromSTEM). Sci Rep, 2022. 12(1): p. 12198.

43. Virk, R.K.A., et al., Disordered chromatin packing regulates phenotypic plasticity. Sci Adv, 2020. 6(2): p. eaax6232.

44. Gladstein, S., et al., Measuring Nanoscale Chromatin Heterogeneity with Partial Wave Spectroscopic Microscopy. Methods Mol Biol, 2018. 1745: p. 337–360.

45. Almassalha, L.M., et al., Macrogenomic engineering via modulation of the scaling of chromatin packing density. Nat Biomed Eng, 2017. 1(11): p. 902–913.

46. Gould, T.J., et al., Defining the epichromatin epitope. Nucleus, 2017. 8(6): p. 625–640.

47. Subramanian, H., et al., Optical methodology for detecting histologically unapparent nanoscale consequences of genetic alterations in biological cells. Proc Natl Acad Sci U S A, 2008. 105(51): p. 20118–23.

48. Subramanian, H., et al., Nanoscale Cellular Changes in Field Carcinogenesis Detected by Partial Wave Spectroscopy. Cancer Research, 2009. 69(13): p. 5357–5363.

49. Eid, A.e.a., Characterizing chromatin packing scaling in whole nuclei using interferometric microscopy. Opt. Lett., 2020. 45: p. 4810–4813.

50. Backman, V. and H.K. Roy, Advances in biophotonics detection of field carcinogenesis for colon cancer risk stratification. J Cancer, 2013. 4(3): p. 251–61.

